# Dose of approved COVID-19 vaccines is based on weak evidence: a review of early-phase, dose-finding trials

**DOI:** 10.1101/2022.09.20.22276701

**Authors:** David Dunn, Richard Gilson, Sheena McCormack, Laura McCoy

**Author notes:** Correspondence to: Professor David Dunn, MRC Clinical Trials Unit at UCL, 90 High Holborn, London WC1V 6LJ.

## Abstract

Although over 12 billion COVID-19 vaccine doses have been administered globally, the important issue of whether the optimal doses are being used has been relatively neglected. To address this question, we reviewed the reports of early-phase dose-finding trials of the nine COVID-19 vaccines approved by World Health Organization (and one additional vaccine which showed partial clinical efficacy), extracting information on study design and findings on reactogenicity and early humoral immune response. The number of different doses evaluated per vaccine varied widely (range 1-7), as did the number of subjects studied per dose (range 15-190). As expected, the frequency and severity of adverse reactions generally increased at higher doses, although most were clinically tolerable. Higher doses also tended to elicit better immune responses, but differences between the maximum dose and the second-highest dose evaluated were small, typically less than 1.6-fold for both binding antibody concentration and neutralising antibody titre. All of the trials had at least one important design limitation: few doses evaluated, large gaps between adjacent doses, or an inadequate sample size. In general, it is therefore uncertain whether the single dose taken into clinical efficacy trials, and subsequently authorised by regulatory agencies, was optimal. In particular, the recommended doses for some vaccines appear to be unnecessarily high. Although reduced dosing for booster injections is an active area of research, the priming dose is equally deserving of study. We conclude by suggesting some ways in which the design of future trials of candidate COVID-19 vaccines could be improved.

## INTRODUCTION

By July 2022, 12.3 billion COVID-19 vaccine doses had been administered globally, with 66.9% of the world population having received at least one dose.(1) However, their distribution had been highly inequitable with the number of doses per 100 people ranging from 27.6 in low-income countries to 202.9 in high-income countries.(1, 2) There is considerable interest in exploring the use of reduced vaccine doses (“fractional” dosing) to stretch the global COVID-19 vaccine supply, lower the cost, and reduce the incidence of adverse reactions.(3, 4) However, the interest in reduced doses interest has mainly focussed on booster vaccine injections rather than the priming dose.(5)

To date, regulatory authorisation for the vaccines has been granted on the basis of the results from large, phase 3 clinical efficacy trials using a COVID-19 endpoint. All of the phase-3 clinical efficacy trials have evaluated a single vaccine dose (compared with placebo) with this dose being informed by preceding early-phase dose-finding studies. Here we examine the design, results, and interpretation of the early-phase dose-finding trials of the approved COVID-19 vaccines. From this we draw conclusions which could lead to improvements in the design of future trials of candidate COVID-19 vaccines and other vaccines in general.

## MATERIALS AND METHODS

We reviewed the early-phase dose-finding trials of the COVID-19 vaccines granted emergency use listing (EUL) by the World Health Organization by the end of July 2022.(6) Eleven vaccines have been approved, although two are different formulations of the same vaccine. In addition, we included the trial of CVnCoV, which narrowly failed to meet the prespecified statistical success criteria for protection against symptomatic disease.(7) The vaccines are listed in Table 1: three mRNA vaccines (BNT162b2, mRNA-1273, CVnCoV), three viral vector vaccines (ChAdOx1, Ad26.COV2.S, Convidecia), one recombinant protein vaccine (NVX-CoV2373), and three whole virus inactivated vaccines (CoronaVac, BBIBP-CorV, BBV152).(8-17) We acknowledge this is not a comprehensive, systematic review; however, with over one hundred COVID-19 vaccines having been evaluated in humans such a review would be less relevant and superficial.(6)

**Table 1.**
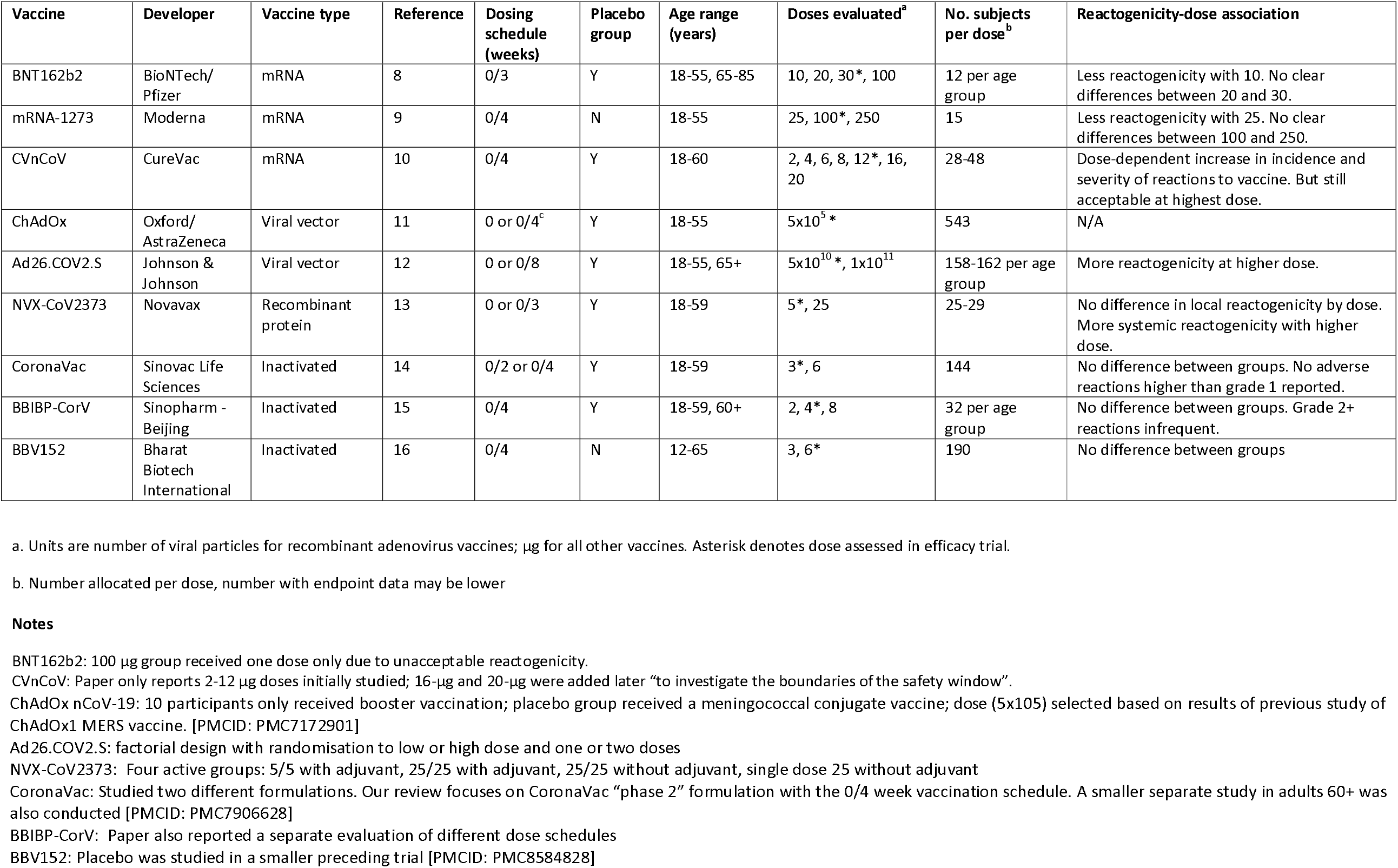
Early-phase dose-finding trials of the licensed COVID-19 vaccines: design aspects and reactogenicity-dose associations.

## RESULTS

### Design features

Table 1 shows the key design features of the reviewed trials. Most assessed a prime-boost strategy, although the Ad26.COV2.S trial employed a factorial design to evaluate the effects of both dose and a single versus two dose regimen. All trials, apart from mRNA-1273 and BBV152, included a placebo group. A recent systematic review found a high rate of reported adverse events in the placebo groups of COVID-19 vaccine trials, suggesting its importance as a baseline, comparator group.(18) Conversely, some vaccine reactogenicity is expected and the key issue is arguably whether the degree of reactogenicity is clinically tolerable. A placebo group is also of limited value in the immunogenicity analyses, apart from providing quality control data and information on the incidence of natural infection in the trial cohort.

The number of different doses evaluated varied widely between the trials. ChAdOx1 assessed a single dose only, partly because standardisation of dose is particularly challenging for viral vector vaccines.(19) Of interest, 24% of subjects in the subsequent efficacy trial inadvertently received a first dose of vaccine that was approximately half that of the planned dose, and an interim analysis reported unexpectedly higher efficacy among these subjects than those who received two standard doses.(20) The trials of inactivated vaccines examined either two or three doses across a relatively narrow dosage range, with a 2-fold difference between adjacent doses. Developers of the more novel mRNA vaccines (BNT162b2, mRNA-1273, CVnCoV) assessed a wider dosage range (10-fold), often with wide gaps between adjacent doses. The number of subjects also varied widely, ranging from 15 per dose (mRNA-1273) and 24 per dose (BNT162b2) to 190 subjects per dose (BBV152). The rationale for the sample size was usually subjective e.g. characterising immune response and/or safety, limited available vaccine, or following national guidelines. Only one trial (BBV152) included a formal statistical power calculation.

All studies reported local and systemic reactions after each vaccination (usually solicited for 7 days), as well as longer-term unsolicited adverse events. However, the reporting of safety outcomes was not standardised across studies. One study recorded whether the second vaccination had been withheld or delayed due to reactogenicity following the first vaccination, which is arguably the most clinically relevant outcome.(14) Serious adverse reactions, such as blood clots and myocarditis, are too rare to be reliably detected in small, early-phase trials.

The timing and details of the immunology assessments are shown in Table 2. Although methodologies were highly variable, subsequent analyses of immune response are made within trials rather than between trials. Most trials quantified the level of binding antibodies against the spike protein (which all the vaccines aimed to induce). The trial of BBIBP-CorV measured neutralising antibodies only, and CoronaVac and Convidecia quantified anti-RBD antibodies. All studies measured neutralising antibodies assessed against Wuhan strains of live or pseudo-virus (or both), although only a subset of participants were tested in the Ad26.COV2.S trial. Few primary publications reported the results of T-cell assays, either because none had been performed or experiments had not been completed in time.

**Table 2.** Details of the immunology assays.

| **Vaccine** | **Timing of immunology assessments (weeks)<sup>a</sup>** | **Binding antibodies** | **Neutralisation antibodies** | **T cell assays** |
| --- | --- | --- | --- | --- |
| BNT162b2 | 0, 1, 3, 4*, 5 | Anti-S1 and anti-RBD IgG (Luminex immunoassay) | Pseudo virus. NT<sub>50</sub>, NT<sub>90</sub>. | Not reported, studies ongoing at time of publication |
| mRNA-1273 | 0, 2, 4, 5, 6, 8* | Anti-S-2P and anti-RBD IgG (ELISA, performed at the NIAID Vaccine Research Center. | Live virus (plaque-reduction neutralization assay), Pseudo virus. NT<sub>50</sub>. Live virus assessed at weeks 0 and 6 only. | Cytokine-staining assay. Data available on 25-µg and 100-µg groups only at time of report. No comparisons reported. |
| CVnCoV | 0, 1, 2, 5, 6*, 8 | Anti-S and anti-RBD IgG (in-house ELISA) | Live virus (microneutralization assay, cytopathic effect read out). NT<sub>50</sub>. | None reported. Analyses of T-cell and B-cell memory responses ongoing at time of publication |
| ChAdOx | 0, 4 | IgG against trimeric spike protein (in-house indirect ELISA).<br>Anti-S and anti-RBD (Meso Scale Discovery multiplexed immunoassay) | Live virus (plaque reduction neutralisation test), Pseudo virus. NT<sub>50</sub>. | Ex-vivo interferon-γ enzyme-linked immunospot (ELISpot) assay |
| Ad26.COV2.S | 0, 2, 4, 8*, 10 | Anti-S IgG (ELISA, developed and qualified at Nexelis, Laval, Canada. | Live virus (microneutralization assay). Random subset of participants (~50 per group). NT<sub>50</sub>, NT<sub>80</sub>. | S-specific T-cell responses measured by cytokine-staining assay at day 15. |
| NVX-CoV2373 | 0, 1, 3, 4, 5* | Anti-S IgG (ELISA, performed at Novavax Clinical Immune Laboratory, Gaithersburg, MD). | Live virus (microneutralization assay). NT<sub>99</sub>. | Cytokines measured in small subgroup of subjects. Numbers insufficient to allow comparison between doses. |
| CoronaVac | 0, 4, 5, 6, 8* | RBD-specific IgG (in-house ELISA from Sinovac) | Live virus (micro cytopathogenic effect assay). Percent neutralisation not specified. | Not assessed |
| BBIBP-CorV | 0, 1, 2, 4, 6* | Not assessed | Live virus. Percent neutralisation not specified. | Not assessed |
| BBV152 | 0, 4, 6*, 8 | Anti-S (s1), anti-RBD, anti- nucleocapsid IgG (in-house ELISA) | Live virus (microneutralisation and plaque reduction neutralisation assays). NT<sub>50</sub>. | Subset of participants at weeks 6 and 8. Th1 and Th2 cytokines; T-cell memory response (CD4+CD45RO+ and CD4+CD45RO+CD27-). |

a. As reported in paper. Protocols may also specify later assessments. Asterisk denotes time-point shown in Figure 1.

### Safety

A narrative summary of the reactogenicity findings is given in Table 1. The inconsistent way in which these data were recorded and/or reported precluded a systematic quantitative analysis. No association between dose and reactogenicity was observed for the inactivated vaccines, however the frequency and severity of adverse reactions generally increased at higher doses for the other types of vaccine. No dose was found to result in clinically unacceptable reactogenicity, with the exception of the 100-µg dose of BNT162b2, which was abandoned.

### Immune response

Following the final injection, all of the vaccines achieved seroconversion rates (for both binding and neutralising antibodies) equal or close to 100%, apart from CVnCoV (range: 69-95% anti-S IgG, 56-83% neutralising antibodies). For each vaccine, we extracted data at the primary analysis timepoint, generally 2-4 weeks after the final. This information was used to construct Figure 1, which shows the average immune response, relative to the lowest dose, for each vaccine. The raw data are included in the Supplementary Appendix.

**Figure 1.**
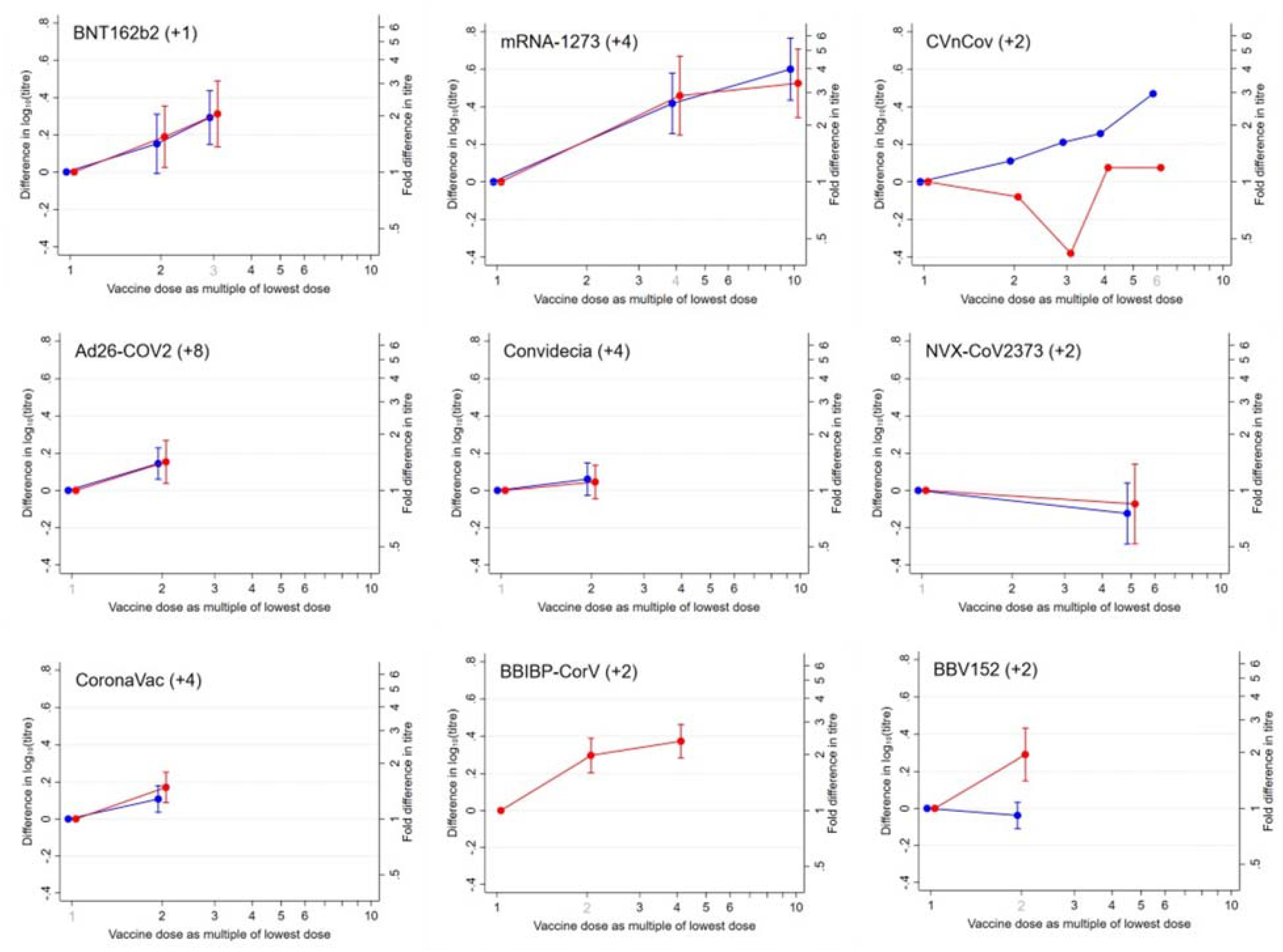
Binding antibody and neutralising antibody response for individual vaccines, relative to the lowest dose <u>Footnote</u> Binding antibodies, blue line; neutralising antibodies, red line. Bars show 95% confidence intervals. Where the confidence interval does not include zero, the difference between that dose and the lowest dose is statistically significant at P<0.05. Dose evaluated in phase-3 efficacy trial shown in grey. Value in brackets denotes number of weeks after final injection when immunology was assessed. BNT162b2: Age groups combined. mRNA-1273: Pseudovirus neutralisation shown. CVnCoV: Values shown are median titres. Confidence intervals not derivable from published data ChAdOx: not included as assessed single dose Ad26.COV2.S: Analysis based on Cohort 1a (18-55 years), where data are more mature. Week 8 timepoint analysed for both single dose and two dose schedules (day of second vaccination for latter group). NT_50_ values shown. Convidecia: anti-RBD binding antibody titres and pseudovirus neutralisation shown. NVX-CoV2373: Two dose group that included adjuvant shown. CoronaVac: “Phase 2” formulation with the 0/4 week vaccination schedule analysed. anti-RBD binding antibody titres shown. BBIBP-CorV: did not assess binding antibodies; age groups combined. BBV152: Plaque-reduction neutralisation assay analysed.

The association between relative response and vaccine dose was very similar for binding and neutralising antibodies, apart from BBV152 (where an association was observed for neutralising antibodies but not binding antibodies), and CVnCoV (where the reverse was observed). There was no evidence of a dose effect for Ad26.COV2.S, Convidecia, or NVX-CoV2373, while CoronaVac showed a weak effect. An approximate linear relationship was observed across the dose range of BNT196b and CVnCoV. For mRNA-1273 and BBIBP-CorV, the relationship was curve-linear, appearing to show a plateau effect.

Immune response also tended to improve at higher doses; however, differences between the maximum dose and the second-highest dose were small, typically less than 0.2 log_10_ (1.6-fold) for both binding antibody concentration and neutralising antibody titre. The clinical interpretation of these differences is hampered by the incomplete understanding of the immunes correlates of protection.(21) However, useful insights were provided by an analysis from the mRNA-1273 efficacy trial, which estimated the hazard ratios of the risk of COVID-19 according to anti-spike IgG and pseudo-virus NT_50_ values measured four weeks after the second vaccination.(22) The authors found that a 0.2 log_10_ lower response in anti-spike IgG concentration predicts a 8.7% (95% CI: 2.6-14.9%) increase in the risk of COVID-19, and a 0.2 log_10_ lower response in NT_50_ predicts a 18.9% (9.0-29.9%) increase. These are modest clinical effects, although the analysis should be interpreted cautiously as follow-up extended to only 16 weeks after the second vaccination, and as findings may not generalise to other vaccines.

The only vaccines for which substantive T-cell data were reported were Ad26.COV2.S, Convidecia, and BBV152. For Ad26.COV2.S and Convidecia, no association between dose and T-cell response was found; for BBV152, a more pronounced T-cell memory response was observed in the higher dose (6-µg) group.

## DISCUSSION

### Dose selected for the phase-3 efficacy trial

Five trials assessed two different doses (Ad26.COV2.S, Convidecia, NVX-CoV2373, BBV152, CoronaVac). Three found a similar immunological effect of the lower dose and higher dose, and the lower dose was selected for the efficacy trial (Ad26.COV2.S, Convidecia, NVX-CoV2373). The higher dose of the BBV152 vaccine elicited a better neutralising antibody response (but a similar binding antibody response) and was taken forward. Finally, the higher dose of the CoronaVac vaccine elicited marginally better responses (differences of log_10_ 0.1-0.2). Pragmatically, the researchers took the lower dose forward on the grounds of production capacity. Three trials assessed three different doses (BNT162b2, mRNA-1273, BBIBP-CorV). The pattern of results was similar for all three vaccines, with the lowest dose being immunologically inferior but no clear difference between the high and intermediate doses. The highest dose was evaluated in the efficacy trials of BNT162b2 and mRNA-1273, while the efficacy trial of BBIBP-CorV evaluated the intermediate dose. Finally, the highest dose (12-µg) of CVnCoV was taken forward for efficacy evaluation, although the researchers acknowledged that the optimal dose could have been higher than this, and the phase-2 trial was extended to examine 16-µg and 20-µg doses after the efficacy trial was initiated.

### Dose selection – general considerations

Although there is a large body of methodological literature on optimal designs for dose-finding studies, the innovations proposed have been rarely used in applied research.(23, 24) Much of this work considers fixed designs (i.e. the doses evaluated and the number of subjects per dose are pre-specified), with the design optimised to find the most accurate estimate of a target dose. The target dose can be defined in various ways – the most relevant for COVID-19 vaccines is arguably the dose achieving a specified, acceptably high fraction of the maximum treatment (known as the ED_P_).(23, 24) Figure 2 illustrates this idea heuristically; this shows a plausible curve-linear relationship between vaccine efficacy and vaccine dose, with an acceptable target dose lying between the dotted lines.(4) As phase-3 efficacy trials generally examine a single dose only, this curve is hypothetical and cannot be validated. Instead, there is an underlying, implicit assumption that the association between immune response (for the primary immunological marker) and dose closely mirrors the association between clinical efficacy and dose.

**Figure 2.**
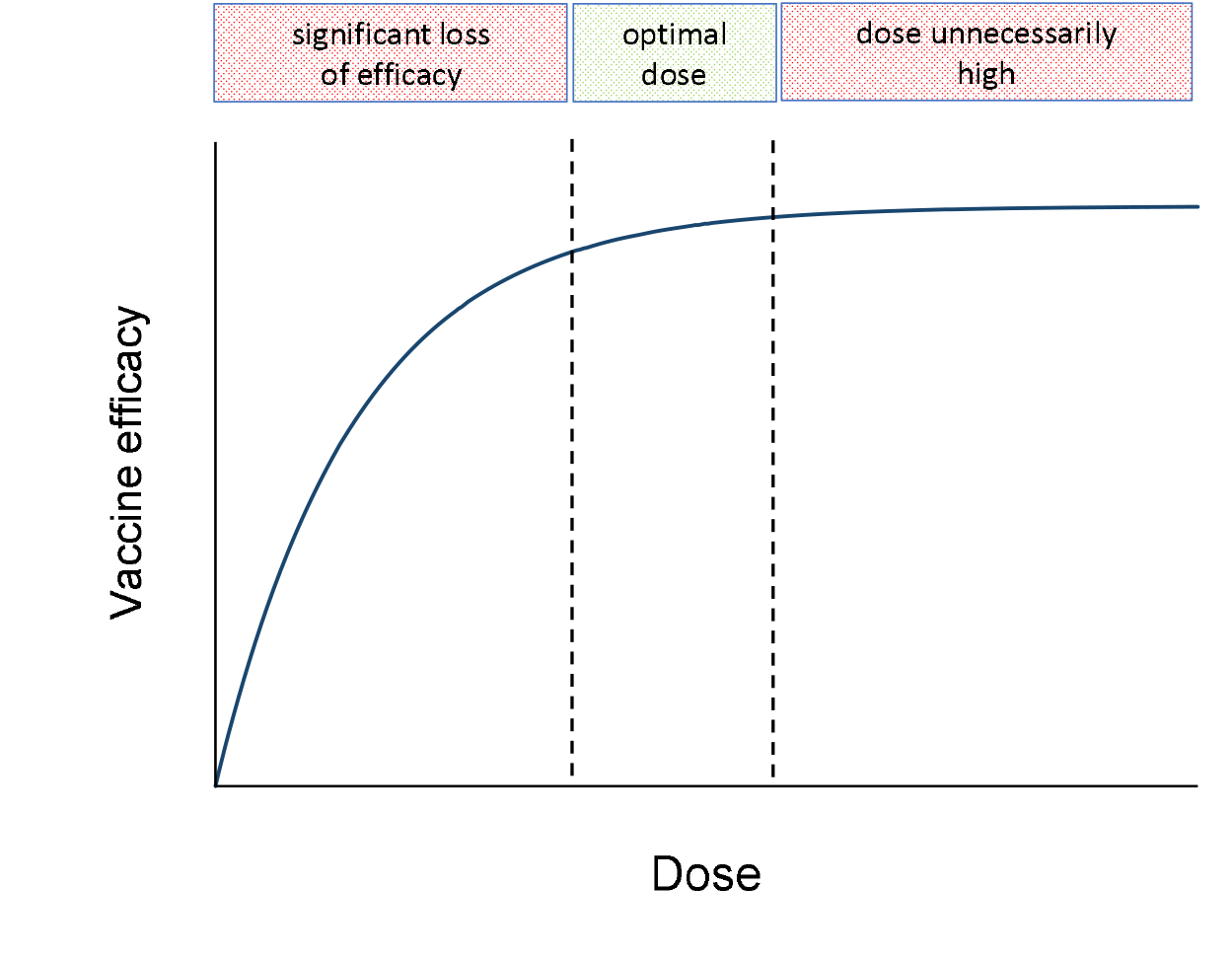
Hypothetical relationship between vaccine efficacy and dose

### Limitations in trial design

All of the trials in this review had at least one important design limitation: few doses evaluated, doses widely dispersed, or an inadequate sample size. First, if only two doses are assessed the best dose cannot possibly be identified – if the low dose elicits a similar immune response to the high dose then an even lower dose may be just as efficacious; if the high dose elicits a better immune response than the low dose then a higher dose may be even more efficacious. Even with three doses, accurate estimation of the dose-response curve is not possible.(24) Second, identification of the optimal dose is compromised if adjacent doses are widely separated. For example, the 100-µg dose of mRNA-1273 out-performed the 25-µg dose but the possibility that an intermediate dose could have been as effective, or almost as effective, as 100-µg cannot be ruled out. Notably, the developers subsequently conducted another phase-2 study that compared 100-µg versus 50-µg and found no difference in reactogenicity or immunogenicity. However, the results of this study were too late to influence the dose used in the phase-3 efficacy trial of this vaccine.(25)

Finally, the precision of the average response value calculated at each dose depends on the sample size. The sample was particularly small in the trials of mRNA-1273 (15 subjects per dose) and BNT162b2 (12 subjects per dose in each of two age groups), giving rise to wide confidence intervals when comparing different doses. This is acknowledged in the BNT162b2 paper: “With 10 to 12 valid results per assay from samples that could be evaluated for each group at each time point, pair-wise comparisons are subject to error and have no clear interpretation”.(8) This implicitly acknowledges that the trial was inadequately powered to identify the optimal dose. Although the developers of these two vaccines could not reasonably have predicted that their vaccines would have been so successful, the mismatch between these sample sizes and the number of doses which have been supplied worldwide (over nine billion) is extremely stark.(26)

### Improving the design of future trials

Our review should not be construed as a criticism of the scientists, working under intense time pressures, who designed and conducted the original dose-finding studies. Also, the regulators had to make pragmatic decisions to ensure safe and effective vaccines were made available as quickly as possible, while acknowledging uncertainties regarding the optimum dose. However, some lessons can be learned which can hopefully improve the design of future studies. The regulatory landscape for COVID-19 vaccines has changed and licensure can now be granted on the basis of neutralising antibody responses compared with approved vaccines.(27) However, this does not avoid the problem of identifying the dose to be included in the licensure application.

A difficult issue in designing COVID-19 dose-finding studies is deciding the range of doses to study, particularly for mRNA vaccines. This is informed by prior dose-ranging studies in animal models, but extrapolation to humans is problematic.(21) Thus, it is prudent to study a wide range of doses, although this means that resources are spread thinly, with a small number of subjects studied per dose. Also, evidence may emerge quickly that some doses are demonstrably too low or too high, as occurred, for example, in the trial of CVnCoV.(10) Adaptive designs mitigate these problems; in these, the dose received by a subject depends on outcomes observed on previous subjects, rather than fixing the doses evaluated and the sample size per dose in advance.(28) Adaptive designs take longer to conduct, and time pressures precluded their use for the first generation of COVID-19 vaccines, but this is now less of a constraint.

## CONCLUSIONS

The use of reduced doses is being actively explored for booster vaccinations and several trials have already reported findings. The largest of these, the COV-BOOST trial, assessed the safety and immunogenicity of seven COVID-19 vaccines as a third dose following two doses of ChAdOx1 or BNT162b2.(29) This included three vaccines which were studied both as a full dose and as a half dose: BNT162b2, NVX-CoV2373, and Valneva (a whole, inactivated virus). The reduced doses of BNT162b2 and NVX-CoV2373 produced potent immune response, with only a minimal decrease in anti-spike IgG and neutralising antibody levels. Also, the FDA have approved a 50-µg half dose of mRNA-1273 when used as a homologous booster injection.(30) This was based on a phase-2 study of 344 participants, in whom the lower dose boosted neutralizing titres significantly above the phase-3 benchmark.(31)

In summary, our review has highlighted the weak evidence base for the licensed doses currently being used in the primary vaccine series. Trials of reduced doses should be widened to include the priming injection as well as booster injections.(3, 4) The experience with COVID-19 vaccines mirrors that in therapeutic drug medicine, where the initially marketed dose is frequently found to be unnecessarily high.(23, 32) The high barrier to achieving a licensure change in dosage highlights the importance of carefully designed dose-finding trials to determine the optimal dose at the earliest opportunity.

## Data Availability

All data produced in the present work are contained in the manuscript

## ACKNOWLEDGEMENTS

DTD and SMc were supported by the UK Medical Research Council (MC_UU_00004/04).

## Notes

### Competing Interest Statement

The authors have declared no competing interest.

### Funding Statement

UK Medical Research Council: salary support

