## Supplementary Appendix for "Dose of approved COVID-19 vaccines is based on weak evidence: a review of early-phase, dose-finding trials"

Table. Data used to generate Figure 1

| **Vaccine** | **Dose** | **Stratum^a^** | **Binding antibody** | | | **Neutralising antibody** | | | **Notes** |
| --- | --- | --- | --- | --- | --- | --- | --- | --- | --- |
|  |  |  | **n** | **GM** | **SE(log_10_GM)** | **n** | **GM** | **SE(log_10_GM)** |  |
| BNT162b2 | 10 | 1 | 12 | 5782 | 0.0897 | 12 | 157 | 0.1026 | Data extracted from Figure 4 in reference 8. SEs were estimated from width of confidence intervals. Stratum 1, ages 18-55; stratum 2, ages 65-85. |
| BNT162b2 | 10 | 2 | 11 | 3203 | 0.0769 | 11 | 79 | 0.1000 |  |
| BNT162b2 | 20 | 1 | 12 | 12464 | 0.0833 | 12 | 363 | 0.0789 |  |
| BNT162b2 | 20 | 2 | 12 | 3056 | 0.1282 | 12 | 84 | 0.1158 |  |
| BNT162b2 | 30 | 1 | 12 | 9136 | 0.0769 | 12 | 361 | 0.0921 |  |
| BNT162b2 | 30 | 2 | 12 | 7985 | 0.1026 | 12 | 149 | 0.1316 |  |
| mRNA-1273 | 25 |  | 13 | 299751 | 0.0830 | 13 | 81 | 0.1016 | Data extracted from Table 2 in reference 9.  Pseudo virus neutralisation analysed |
| mRNA-1273 | 100 |  | 14 | 782719 | 0.0519 | 14 | 232 | 0.0778 |  |
| mRNA-1273 | 250 |  | 13 | 1192154 | 0.0570 | 14 | 270 | 0.0454 |  |
| CVnCoV | 2 |  | 34 | 1738 |  | 34 | 48 |  | Data extracted from Immunogenicity section of reference 10. Values are medians and not geometric means. SEs were not derivable. |
| CVnCoV | 4 |  | 34 | 2239 |  | 34 | 40 |  |  |
| CVnCoV | 6 |  | 36 | 2818 |  | 36 | 20 |  |  |
| CVnCoV | 8 |  | 34 | 3136 |  | 34 | 57 |  |  |
| CVnCoV | 12 |  | 20 | 5118 |  | 34 | 57 |  |  |
| Ad26-COV2 | 5x10^10^ | 1 | 65 | 873 | 0.0469 | 24 | 370 | 0.0677 | Data extracted from Figure 2 in reference 12.  SEs were estimated from width of CIs. Analysis based on Cohort 1a (18-55 years). Stratum 1, single dose schedule; stratum 2, two dose schedule. Week 8 value analysed i.e. day of booster vaccination. NT_50_ nAb values analysed. |
| Ad26-COV2 | 5x10^10^ | 2 | 70 | 660 | 0.0573 | 24 | 310 | 0.0625 |  |
| Ad26-COV2 | 1x10^11^ | 1 | 71 | 1100 | 0.0417 | 24 | 488 | 0.0833 |  |
| Ad26-COV2 | 1x10^11^ | 2 | 74 | 754 | 0.0573 | 25 | 288 | 0.0625 |  |
| Convidecia | 5x10^10^ |  | 129 | 571 | 0.0443 | 129 | 55.3 | 0.0442 | Data extracted from Results section of reference 13. anti-RBD binding antibody titres shown. Pseudo virus neutralisation analysed |
| Convidecia | 1x10^11^ |  | 253 | 656 | 0.0293 | 253 | 61.4 | 0.0324 |  |
| NVX-CoV2373 | 5 |  | 29 | 63160 | 0.0649 | 29 | 3906 | 0.0940 | Data extracted from Tables S10 and S11 in Supplementary Appendix for reference 14. Two dose group that included adjuvant analysed. |
| NVX-CoV2373 | 25 |  | 27 | 47521 | 0.0755 | 27 | 3305 | 0.0897 |  |
| CoronaVac | 3 |  | 117 | 1784 | 0.0356 | 117 | 44 | 0.0375 | Data extracted from Table 6-1 in Supplementary Appendix for reference 15. Phase formulation with day 0/28 schedule. anti-RBD binding antibody titres analysed. |
| CoronaVac | 6 |  | 117 | 2287 | 0.0256 | 118 | 65 | 0.0329 |  |
| BBIBP-CorV | 2 | 1 |  |  |  | 32 | 88 | 0.0668 | Data extracted from Results section (p.49) of reference 16. Stratum 1, ages 18-55; stratum 2, ages 65+. Study did not assess binding antibodies. |
| BBIBP-CorV | 2 | 2 |  |  |  | 31 | 81 | 0.0466 |  |
| BBIBP-CorV | 4 | 1 |  |  |  | 32 | 211 | 0.0630 |  |
| BBIBP-CorV | 4 | 2 |  |  |  | 32 | 132 | 0.0431 |  |
| BBIBP-CorV | 8 | 1 |  |  |  | 32 | 229 | 0.0457 |  |
| BBIBP-CorV | 8 | 2 |  |  |  | 30 | 171 | 0.0555 |  |
| BBV152 | 3 |  | 184 | 10414 | 0.0289 | 184 | 101 | 0.0682 | Data extracted from Results section of reference 15. Plaque reduction neutralisation assays analysed |
| BBV152 | 6 |  | 177 | 9542 | 0.0323 | 177 | 197 | 0.0518 |  |

**Notes on analysis**

All analyses were performed on a log_10_ scale. Standard errors were derived by dividing the width of the 95% confidence interval by 3.92 (2x1.96). For studies with more than one stratum, data were averaged (weighted average, by sample size) across the different strata.
